# Transient reduction of sex-specific serum fibrillin-1 concentrations in acute spontaneous cervical artery dissection: a prospective multicenter study

**DOI:** 10.64898/2026.08.20.26360968

**Authors:** Silke Zimmermann, Markus Weißenfels, Norma Krümmer, Wolfgang Härtig, Gesa Weise, Johann Otto Pelz

## Abstract

**Background:** Spontaneous cervical artery dissection (sCeAD) is a rare vasculopathy whose pathophysiology remains incompletely understood. Impaired vascular extracellular matrix integrity, including elastic fibers, may contribute to its development. We investigated whether serum fibrillin-1 and soluble elastin fragments (sELF) differ between patients with sCeAD and controls during the acute and chronic stages.

**Methods:** Patients with acute sCeAD were prospectively enrolled at four German stroke centers. Blood samples were collected at baseline and after 6±1 months. Patients with a first acute ischemic stroke unrelated to sCeAD and healthy individuals served as controls. Serum fibrillin-1 and sELF concentrations were measured using enzyme-linked immunosorbent assays.

**Results:** 61 patients with sCeAD, 53 patients with first non-CeAD ischemic stroke, and 79 healthy controls were included. After sex-matching, serum fibrillin-1 concentrations were significantly lower in patients with acute sCeAD than in healthy controls (97 [60; 192] vs. 176 [113; 269] ng/mL; p=0.009). Fibrillin-1 concentrations were also lower in both male and female patients with sCeAD than in respective healthy controls. In patients with sCeAD, fibrillin-1 concentrations increased significantly after 6 months compared with baseline (171 [130; 270] vs. 104 [67; 205] ng/mL; p=0.021). Serum fibrillin-1 concentrations were higher in men than in women across all study groups. No significant differences in sELF concentrations were observed between groups or time points.

**Discussion:** Serum fibrillin-1 concentrations were lower during acute sCeAD and increased significantly during follow-up, whereas sELF concentrations remained unchanged. These findings support an association between circulating fibrillin-1 and acute sCeAD and warrant further investigation of its role in sCeAD pathophysiology. Pronounced sex-related differences in fibrillin-1 concentrations highlight the importance of sex-specific analyses in future.

## Introduction

Spontaneous cervical artery dissection (sCeAD) is a rare vasculopathy characterized by a non-traumatic disruption of the medial layer of the arterial wall of the internal carotid artery (ICA) or vertebral artery (VA), resulting in an intramural hematoma (Debette 2014). Its annual incidence is approximately 4.6 cases per 100,000 individuals (Shu et al., 2025). With a peak incidence in the fifth decade of life, sCeAD is an important cause of ischemic stroke in younger adults (Ferro et al., 2010).

Despite extensive research, the pathophysiology of sCeAD remains incompletely understood. Increasing evidence suggests that patients with sCeAD have an underlying, often clinically inapparent abnormality of the vascular connective tissue (Gunduz et al., 2023). Disruption of elastic fibers within the arterial wall could potentially lead to the release of extracellular matrix components into the circulation. However, the extent to which circulating extracellular matrix proteins reflect the vascular changes associated with acute sCeAD remains unclear.

Zhu and colleagues previously reported increased serum fibrillin-1 concentrations in patients with acute sCeAD compared with control individuals and patients with non-sCeAD ischemic stroke and proposed fibrillin-1 as a potential biomarker for the diagnosis of sCeAD (Zhu et al., 2018).

Fibrillin-1 is an important component of extracellular microfibrils and contributes to the structural integrity and elasticity of the arterial wall (Schrenk et al., 2018; Li et al., 2024). Fibrillin-1-containing microfibrils provide a scaffold for elastin deposition and are involved in the regulation of elastic fiber homeostasis. Fibrillin-1 also plays an important role in cell–cell and cell–matrix interactions under physiological and pathological conditions. Accordingly, dysregulation of fibrillin-1 has been implicated in a range of human diseases, including cardiovascular disorders (Li et al., 2024). However, only a limited number of studies have investigated circulating fibrillin-1 concentrations in patients with vascular disease (Zhu et al., 2018; Hui et al., 2020).

In the present study, we investigated whether serum fibrillin-1 and soluble elastin fragments (sELF), the latter serving as a circulating surrogate marker of elastin degradation, were altered in patients with sCeAD during the acute and chronic stages of the disease.

## Methods

### Study design and ethics

This multicenter, prospective, non-interventional, exploratory study was conducted in accordance with the ethical standards of the Declaration of Helsinki and its subsequent amendments. The study was approved by the Ethics Committee of the Medical Faculty of Leipzig University, Leipzig, Germany (reference number 410/18-ek). All patients or their legal representatives provided written informed consent.

### Study population

The present study forms part of a prospective research project investigating vascular extracellular matrix components and autoimmune phenomena in patients with sCeAD (Zimmermann et al., 2024; Zimmermann et al., 2025). The study design and patient cohort have been described previously.

Briefly, patients with sCeAD were prospectively enrolled at four German stroke centers between May 2018 and June 2023. The diagnosis of sCeAD involving the ICA or VA was based on typical clinical features, including Horner syndrome, neck pain, headache, or ischemic stroke, together with at least one of the following radiological findings: (1) an intramural hematoma on magnetic resonance imaging; (2) a long, tapering stenosis distal to the carotid bifurcation without evidence of atherosclerosis on computed tomography angiography or magnetic resonance angiography; or (3) an intimal flap or double lumen (Debette et al., 2021). In the presence of significant head or neck trauma, CeAD was classified as traumatic; in the absence of significant trauma, it was classified as spontaneous. sCeAD was considered acute when the first symptoms attributable to the dissection had occurred within 14 days before study enrollment.

Two different cohorts were selected for comparisons: consecutive patients with acute ischemic stroke not attributable to CeAD (non-CeAD ischemic stroke) and healthy individuals without known cerebrovascular or cardiovascular disease.

In patients with sCeAD, blood samples were collected by venipuncture within 48 hours after hospital admission at study enrollment and again after 6 ± 1 months. In patients with non-CeAD ischemic stroke, blood samples were collected within 72 hours after symptom onset.

Blood was collected into serum tubes (S-Monovette®, Sarstedt AG & Co., Nümbrecht, Germany). After collection, tubes were kept upright for 30 minutes to allow clot formation and were subsequently centrifuged at 5,500 × g for 10 minutes. Serum aliquots were stored at −80 °C until analysis.

### Measurement of fibrillin-1 and soluble elastin fragments

All baseline and follow-up samples were analyzed in the same analytical period. Serum concentrations of fibrillin-1 and sELF were determined using quantitative sandwich enzyme-linked immunosorbent assays (ELISAs) according to the manufacturers’ instructions. Fibrillin-1 was measured using an ELISA from Invitrogen by Thermo Fisher Scientific (Waltham, MA, USA), whereas sELF was measured using an ELISA from MyBioSource (San Diego, CA, USA).

Samples were diluted according to the respective assay ranges using the dilution buffer provided by the manufacturers. Assay precision was assessed by the intra-assay and inter-assay coefficients of variation. For fibrillin-1, intra- and inter-assay coefficients of variation were both <7%, while for sELF, intra- and inter-assay coefficients of variation were <8% and <10%, respectively, indicating good analytical precision.

Briefly, standards and samples were added to antibody-coated wells, allowing fibrillin-1 or sELF to bind to the immobilized capture antibodies. After removal of unbound material, biotin-conjugated detection antibodies were added, followed by avidin-conjugated horseradish peroxidase. After washing, substrate solution was added and color development was measured. Signal intensity was proportional to the amount of antigen present in the sample.

### Statistical analysis

Statistical analyses were performed using IBM SPSS Statistics, version 29 (IBM Corp., Armonk, NY, USA). Continuous variables were summarized as mean ± standard deviation and, because of extreme outliers, as median with first and third quartiles. Categorical variables were expressed as absolute numbers and percentages.

Categorical variables were compared using the chi-square test. The Mann–Whitney U test was used for comparisons between independent groups, and the Wilcoxon signed-rank test was used for paired observations. The Kruskal–Wallis H test was used as a global non-parametric test for independent groups, and the Friedman test was used for dependent groups.

Given the significant differences in demographic characteristics between study groups, multivariable linear regression analyses were performed to determine the independent associations of age, biological sex, and study group with serum fibrillin-1 and sELF concentrations. For demographic factors showing a significant association with biomarker concentrations, case-control matching was subsequently performed to account for potential confounding. Biomarker concentrations were then compared between matched cases and controls using the Wilcoxon signed-rank test.

A two-sided p value <0.05 was considered statistically significant.

## Results

### Study population

A total of 76 patients with CeAD were initially enrolled. Eleven patients (14.5%) were subsequently excluded: seven because CeAD could not be confirmed after extensive diagnostic evaluation and four because symptom onset had occurred more than 14 days before study enrollment (Figure 1).

**Figure 1:**
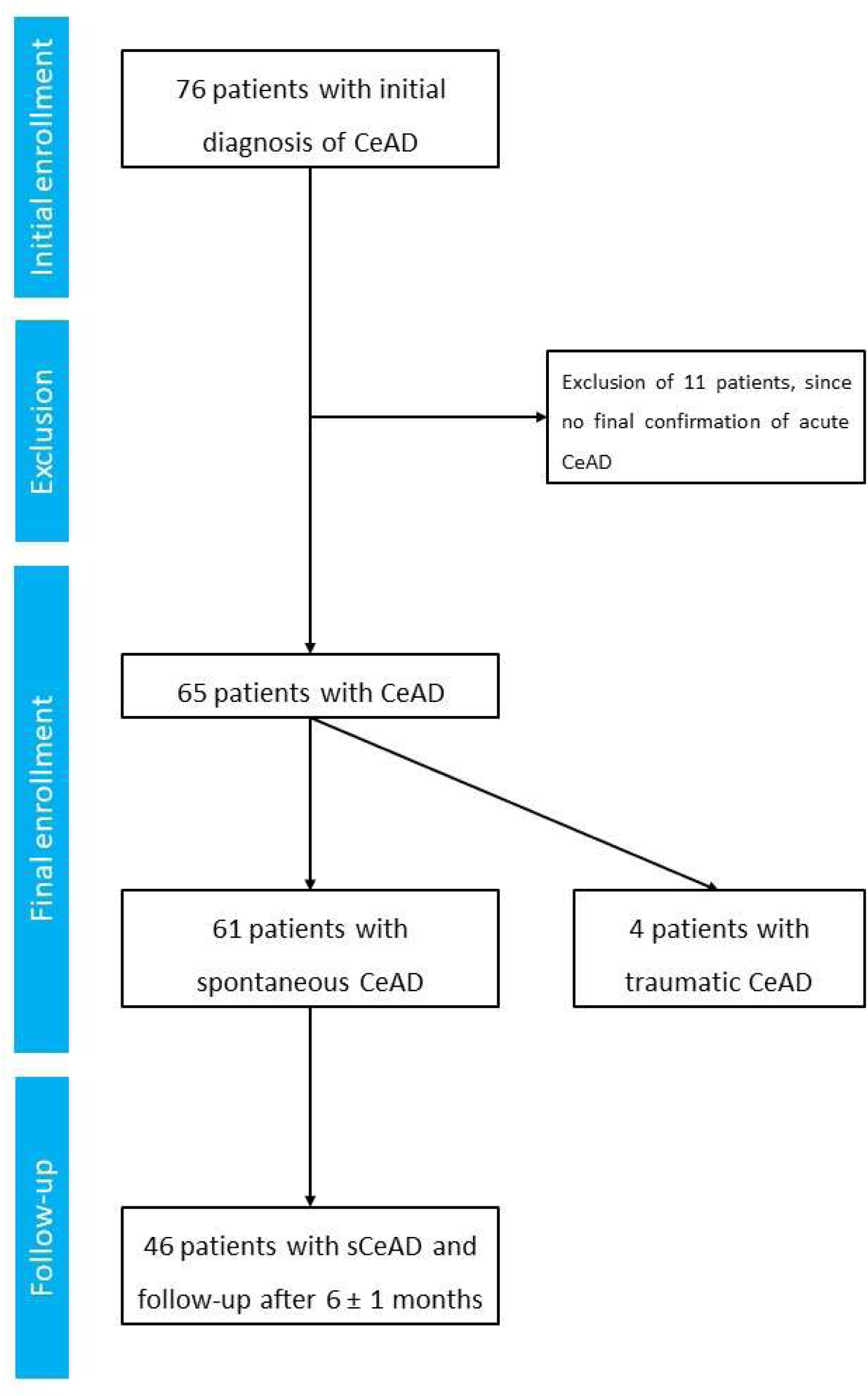
Flow chart of study participant enrollment. The flow chart shows the enrollment, exclusion, and allocation of patients with spontaneous cervical artery dissection (sCeAD).

Of the remaining 65 patients, 61 had spontaneous and four had traumatic CeAD. The 61 patients with sCeAD had a mean age of 45.6 ± 10.1 years, and 21 (34.4%) were women. Fifteen patients with sCeAD were lost to follow-up, resulting in 46 (75.4%) patients with blood collection after 6 ± 1 months. The control cohorts comprised 53 patients with first acute ischemic stroke unrelated to CeAD (mean age, 56.4 ± 13.8 years; 15 women (28.3%)) and 79 healthy controls (mean age, 57.6 ± 12.9 years; 55 women (69.6%)). Baseline characteristics are summarized in Table 1.

**Table 1:** Baseline demographic and clinical characteristics of the study population. *P values refer to comparisons across the three main study groups (acute sCeAD, non-CeAD ischemic stroke, and healthy controls); traumatic CeAD was not included in inferential statistical analyses because of the small sample size*. sCeAD, spontaneous cervical artery dissection; SD, standard deviation; ICA, internal carotid artery; VA, vertebral artery.

|  | Patients with acute sCeAD<br>(n = 61) | Patients with<br>traumatic CAD (n = 4) | Patients with non-CeAD<br>ischemic stroke (n = 53) | Controls<br>(n = 79) | p value |
| --- | --- | --- | --- | --- | --- |
| Age in years<br>(mean ± SD) | 45.6 ± 10.1 | 54.8 ± 4.2 | 56.4 ± 13.8 | 57.6 ± 12.9 | < 0.001 |
| Female sex<br>(n, %) | 21 (34.4 %) | 1 (25.0 %) | 15 (28.3 %) | 55 (69.6 %) | <0.001 |
| Location of CAD<br>(n, %) | ICA: 39 (63.9 %)<br>VA: 22 (36.1 %) | ICA: 3 (75.0 %)<br>VA: 1 (25.0 %) | - | - | - |
| Arterial Hypertension<br>(n, %) | 31 (50.8 %) | 3 (75.0 %) | 38 (71.7 %) | 29 (36.7 %) | <0.001 |
| Hyperlipidemia<br>(n, %) | 30 (49.2 %) | 0 | 39 (73.6 %) | 17 (21.5 %) | <0.001 |
| Diabetes mellitus<br>(n, %) | 2 (3.3 %) | 1 (25 %) | 12 (22.6 %) | 1 (1.3 %) | <0.001 |
| Current smoking<br>(n, %) | 22 (36.1 %) | 0 | 29 (54.7 %) | 15 (19.0 %) | <0.001 |

The study groups differed significantly with respect to age and biological sex. Patients with sCeAD were younger than individuals in both control groups, whereas the healthy control group contained a substantially higher proportion of women. Because only four patients had traumatic CAD, these patients are presented descriptively but were not included in inferential statistical analyses.

### Serum levels of soluble elastin fragments and fibrillin-1

Due to assay-related reasons, data on fibrillin-1 was available for n = 55 and on sELF for n = 59 patients at baseline, and for n = 44 (fibrillin-1) and n = 46 (sELF), respectively, at 6 ± 1 months. This was mainly due to values below the detection rate, since the visual inspection of the respective wells showed no or just a weak staining.

At baseline, serum fibrillin-1 and sELF concentrations did not differ significantly among patients with sCeAD, patients with non-CeAD ischemic stroke, and healthy controls in the unadjusted global analyses (Kruskal–Wallis test, p = 0.130 and p = 0.101, respectively; Table 2). No significant differences in fibrillin-1 or sELF concentrations were observed between patients with sCeAD of the ICA and VA (Table 3).

**Table 2:** Serum concentrations of serum fibrillin-1 and soluble elastin fragments (sELF) in patients with spontaneous cervical artery dissection (sCeAD) and control groups. Values are presented as median [first quartile; third quartile]. *P values were calculated using the Kruskal–Wallis test across patients with acute sCeAD at baseline, patients with non-CeAD ischemic stroke, and healthy controls. The traumatic CeAD group was not included in inferential statistical analyses*.

|  | Patients with<br>sCeAD at baseline | Patients with<br>sCeAD at 6 ± 1<br>months | Patients with<br>traumatic CeAD | Patients with non-sCeAD acute<br>ischemic stroke | Controls | p value |
| --- | --- | --- | --- | --- | --- | --- |
| Fibrillin-1 in ng/ml | (n = 55)<br>104 [67; 205] | (n = 44)<br>171 [130; 270] | (n = 4)<br>100 [58; 130] | (n = 51)<br>138 [92; 178] | (n = 76)<br>147 [101; 243] | 0.130* |
| sELF in ng/ml | (n = 59)<br>58.0 [42.6; 89.4] | (n = 46)<br>64.3 [42.4; 79.7] | (n = 4)<br>58.2 [28.7; 96.6] | (n = 53)<br>58.3 [37.6; 107.5] | (n = 78)<br>64.7 [44.2; 92.5] | 0.101* |

**Table 3:** Serum concentrations of fibrillin-1 and soluble elastin fragments (sELF) according to dissection location and biological sex. Values are presented as median [first quartile; third quartile]. *P values for ICA versus VA comparisons were calculated using the Mann–Whitney U test. Sex-specific comparisons were performed using the Mann–Whitney U test*. ICA, internal carotid artery; VA, vertebral artery.

|  | Patients with sCeAD at baseline |  |  | Patients with sCeAD at baseline |  |  | Patients with non-sCeAD<br>acute ischemic stroke |  |  | Controls |  |  |
| --- | --- | --- | --- | --- | --- | --- | --- | --- | --- | --- | --- | --- |
|  | ICA | VA | p value | male | female | p value | male | female | p value | male | female | p value |
| Fibrillin-1 in ng/ml | (n = 35)<br>104 [67; 196] | (n = 20)<br>105 [68; 259] | 0.594 | (n = 37)<br>169 [85; 263] | (n = 18)<br>84 [58; 109] | **0.006** | (n = 37)<br>149 [107; 224] | (n = 14)<br>96 [76; 96] | **0.011** | (n = 24)<br>243 [149; 405] | (n = 52)<br>127 [88; 172] | **<0.001** |
| sELF in ng/ml | (n = 38)<br>58 [39.7; 107] | (n = 21)<br>56.8 [45.2; 75.9] | 0.869 | (n = 39)<br>54.1 [42; 77] | (n = 20)<br>73.3 [40.5; 131] | 0.158 | (n = 37)<br>59.8 [37.9; 99.9] | (n = 15)<br>55.4 [31.2; 163] | 0.441 | (n = 24)<br>54.0 [39.5;<br>76.5] | (n = 54)<br>70.4 [49.5;<br>96.0] | **0.011** |

Because of the significant differences in age and biological sex between the study groups, multivariable linear regression analyses were performed with serum fibrillin-1 or sELF concentrations as the dependent variable and study group, biological sex, and age as independent variables. Biological sex was significantly associated with serum fibrillin-1 concentrations (β = 0.342, p < 0.001), whereas no significant effects were observed for sELF. To further account for the effect of biological sex on fibrillin-1 concentrations, patients with sCeAD were matched 1:1 to healthy controls according to biological sex. After adjustment for biological sex, serum fibrillin-1 concentrations were significantly lower in patients with acute sCeAD than in healthy controls (97 ng/mL [60; 192] vs. 176 ng/mL [113; 269]; Wilcoxon signed-rank test, p = 0.009).

In sex-stratified analyses, both male and female patients with sCeAD had significantly lower serum fibrillin-1 concentrations than their respective healthy controls (p = 0.017 and p = 0.003, respectively).

Serum fibrillin-1 concentrations were consistently higher in men than in women across all study groups. Among patients with sCeAD, median fibrillin-1 concentrations were 169 ng/mL [85; 263] in men and 84 ng/mL [58; 109] in women (p = 0.006). Corresponding concentrations were 149 ng/mL [107; 224] and 96 ng/mL [76; 96] in male and female patients with non-CeAD ischemic stroke (p = 0.011), respectively, and 243 ng/mL [149; 405] and 127 ng/mL [88; 172] in male and female healthy controls (p < 0.001), respectively (Table 3). Serum sELF concentrations did not differ significantly between men and women with sCeAD or between men and women with non-CeAD ischemic stroke. Among healthy controls, however, sELF concentrations were significantly higher in women than in men (70.4 ng/mL [49.5; 96.0] vs. 54.0 ng/mL [39.5; 76.5]; p = 0.011).

In patients with sCeAD, serum fibrillin-1 concentrations were significantly higher after 6 months than at baseline (171 ng/mL [130–270] vs. 104 ng/mL [67–205]; Wilcoxon signed-rank test, p = 0.021; Figure 2). No significant difference was observed for serum sELF concentrations over the same period (Wilcoxon signed-rank test, p = 0.567).

**Figure 2:**
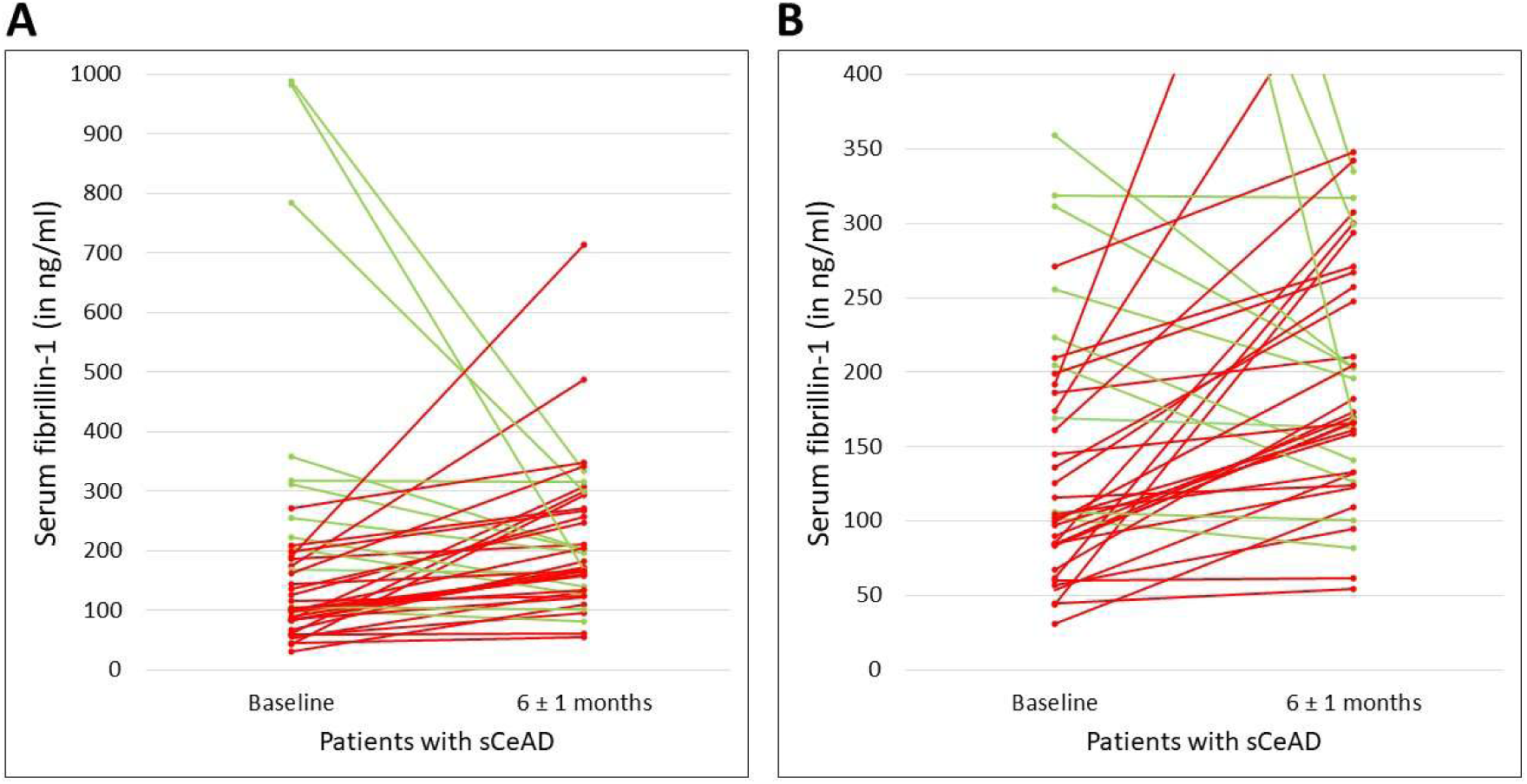
Longitudinal changes in serum fibrillin-1 concentrations in patients with spontaneous cervical artery dissection (sCeAD). (A) Individual serum fibrillin-1 concentrations at baseline and after 6 ± 1 months in patients with spontaneous cervical artery dissection. (B) The same individual patient data are shown with the y-axis truncated at 400 ng/mL to facilitate visualization of the majority of individual trajectories. Each line represents an individual patient. Serum fibrillin-1 concentrations increased significantly from baseline to 6 ± 1 months (Wilcoxon signed-rank test, p = 0.021).

At 6-month follow-up, fibrillin-1 concentrations remained significantly higher in male than in female patients with sCeAD (204 ng/mL [161–299] vs. 131 ng/mL [98–173]; Mann–Whitney U test, p = 0.015).

## Discussion

The main findings of this study are that serum fibrillin-1 concentrations were significantly lower in patients with acute sCeAD than in healthy controls after accounting for biological sex, and that fibrillin-1 concentrations increased significantly during the 6-month follow-up period. Additionally, serum fibrillin-1 concentrations were consistently higher in men than in women across all study groups. In contrast, serum sELF concentrations did not differ significantly between the main study groups or between baseline and follow-up.

Over the past decade, sCeAD has increasingly been considered a multifactorial disorder resulting from an interplay between an underlying, potentially clinically inapparent arteriopathy, environmental risk factors, and transient triggers (Debette 2014; Debette et al., 2014; Gunduz et al., 2023). Several observations support the concept of an underlying vascular connective tissue abnormality. At the macroanatomical level, patients with sCeAD have been reported to have a larger aortic root diameter (Tzourio et al., 1997), increased carotid arterial stiffness (Calvet et al., 2004), impaired vasodilation (Baumgartner et al., 2007), and increased internal carotid artery tortuosity (Mayer-Suess et al., 2025).

Ultrastructural studies have also provided evidence of vascular tissue abnormalities in patients with sCeAD. Examinations of the superficial temporal artery revealed signs of tissue weakening in patients with sCeAD compared with healthy controls (Völker et al., 2005). Furthermore, electron microscopy of skin and arterial tissue from patients with sCeAD demonstrated abnormalities, including elastic fiber fragmentation and medial degeneration (Brandt et al., 2001; Brandt et al., 2005). Mayer-Suess and colleagues previously identified an extracellular matrix protein signature associated with collagen and elastin in patients with recurrent sCeAD, but not in patients with a first single-vessel sCeAD (Mayer-Suess et al., 2020).

Only a limited number of studies have investigated circulating components of the vascular extracellular matrix in patients with sCeAD. In the present study, we focused on fibrillin-1 and sELF because both are associated with the elastic fiber system. We found that serum fibrillin-1 concentrations were significantly higher in men than in women across all study groups and, after accounting for biological sex, were significantly lower in patients with acute sCeAD than in healthy controls. Subsequently, fibrillin-1 concentrations increased during the 6-month follow-up period.

Sex-specific differences have previously been described in the epidemiology of sCeAD, with men being affected more frequently and women tending to be younger at the time of dissection (Arnold et al., 2006; Metso et al., 2012). Sex-related differences have also been reported for connective tissue disorders involving fibrillin-1. Marfan syndrome is a multisystem genetic disorder caused by pathogenic variants in the fibrillin-1 gene (FBN1) and characterized by abnormalities of the connective tissue. In a mouse model of severe Marfan syndrome, male mice showed more rapid aortic dilatation, greater stiffening, and more pronounced extracellular matrix remodeling than female mice (Dwivedi et al., 2024). Furthermore, arterial events in patients with Marfan syndrome and other inherited connective tissue disorders have been reported to occur predominantly in men (Calderon-Martinez et al., 2025). Although these observations do not establish a mechanistic relationship with sCeAD, they support the importance of considering biological sex when investigating fibrillin-1 and vascular connective tissue disorders.

Our findings differ from those reported by Zhu and colleagues, who observed increased serum fibrillin-1 concentrations in patients with acute sCeAD (Zhu et al., 2018). In the present study, fibrillin-1 concentrations were lower in both men and women with acute sCeAD than in healthy controls and subsequently increased during follow-up. Several methodological and biological factors could potentially contribute to these discrepant findings. First, the two studies used different fibrillin-1 ELISA assays, which may differ with respect to antibody specificity, epitope recognition, calibration, and analytical characteristics. Second, the study populations differed geographically and potentially with respect to genetic and other biological characteristics. Interestingly, patients of African ancestry had a higher frequency of the rs9349379 polymorphism of the PHACTR1 gene, a sCeAD risk allele, compared to patients of European ancestry (Debette et al., 2015; Green et al., 2018). However, the available data do not allow us to determine whether population differences contributed to the discrepant results. Another explanation that warrants experimental investigation is analytical interference caused by circulating anti-fibrillin-1 antibodies during the acute phase of sCeAD. Such antibodies could potentially affect immunoassay measurements and result in apparently reduced fibrillin-1 concentrations. This hypothesis was not investigated in the present study; therefore, it remains speculative. Future studies using independent analytical approaches and direct assessment of anti-fibrillin-1 antibodies will be required to test this possibility.

Addressing other circulating extracellular matrix markers, lower serum levels of elastin and collagen type III were reported for the acute phase of sCeAD (Zimmermann et al., 2025). However, elastin is extremely hydrophobic, which raised concerns regarding the reliability of its direct measurement in the serum. Thus, for the current study, we measured sELF, a marker of elastin degradation. Serum sELF concentrations did not differ significantly between the main study groups or over time. In contrast, several studies in patients with acute aortic dissection have reported elevated circulating sELF concentrations and suggested a potential role as a biomarker of acute vascular injury (Shinohara et al., 2002; Peng et al., 2015; Meng et al., 2018). Although both aortic dissection and sCeAD involve the arterial tunica media, the thoracic aorta has a substantially larger diameter and contains more elastic fibers than the distal ICA or VA, which are predominantly affected in sCeAD. The vascular injury associated with sCeAD may therefore be insufficient to produce a measurable systemic increase in circulating sELF. Another possible explanation is the difficult determination of the exact onset of sCeAD. Because the onset of sCeAD may be clinically subtle or initially asymptomatic (Adham et al., 2021), circulating concentrations of extracellular matrix degradation products may already have changed by the time of clinical presentation and blood sampling. Thus, a transient increase in sELF could potentially have occurred before study enrollment and may have been missed by the present sampling strategy. This possibility remains speculative.

Limitations

Despite its strength of being a prospective and multicenter study, this study has several limitations. First, only four patients with traumatic CAD were included, precluding meaningful statistical analysis of this subgroup. Second, healthy controls were selected based on self-reported absence of cerebrovascular and cardiovascular disease. Because no systematic vascular imaging or other vascular assessment was performed, clinically asymptomatic vascular abnormalities cannot be completely excluded. Third, the study groups were not balanced with respect to age and biological sex. Although statistical methods were used to account for these differences, residual confounding cannot be excluded, particularly given the relatively small sample size. The pronounced sex-related differences in serum fibrillin-1 emphasize the importance of appropriately balanced cohorts and prespecified sex-specific analyses in future studies. Finally, the present study was exploratory and does not establish whether altered serum fibrillin-1 concentrations are causally involved in sCeAD or merely reflect processes associated with acute vascular injury. Independent validation in larger cohorts and studies using complementary analytical methods will therefore be required.

## Conclusion

Serum fibrillin-1 concentrations were significantly lower in patients with acute sCeAD than in healthy controls after accounting for biological sex and increased significantly during the subsequent 6-month follow-up period. In contrast, serum sELF concentrations did not differ significantly between study groups or over time. These findings support an association between circulating fibrillin-1 and acute sCeAD but do not establish a causal role. The marked sex-related differences in serum fibrillin-1 also highlight the importance of sex-specific analyses in future studies of sCeAD and other vascular connective tissue disorders.

## Declarations

This manuscript complies with all instructions to authors.

All authorship requirements have been met and the manuscript was approved by all authors for publication.

This manuscript has not been published elsewhere and is not under consideration by another journal.

Ethics approval

This prospective, longitudinal, multicenter study was performed according to the ethical standards laid down in the 1964 Declaration of Helsinki and its later amendments. The study was approved by the local ethics committee of the Medical Faculty at the University of Leipzig, Leipzig, Germany (reference number 410/18-ek). All study participants gave their written informed consent.

Full data access and statements

Johann Otto Pelz as the corresponding author of the manuscript and the principal investigator of the study has full access to the underlying data, and takes full responsibility for the data, the analyses and interpretation, and the conduct of the research.

Availability of data and materials

The dataset underlying this study is available from the corresponding author on reasonable request for any qualified investigator.

## Author contributions

SZ: measurements, analysis of data; MW, NK, and GW: acquisition of data, critical revision of the manuscript; WH: study support and critical revision of the manuscript for intellectual content; JOP: study design, study supervision, data analysis and interpretation, writing the first draft of the manuscript.

## Acknowledgement

The authors thank Susann Kostmann for technical assistance and Rita Lachmund for organizational support.

